# Phantom Fairness: A Reproducible Audit of How Automatically Extracted Labels Can Conceal Demographic Disparities in Chest Radiograph Classifiers

**DOI:** 10.64898/2026.10.03.26364646

**Authors:** Krutarth Patel, Phanindra Beedala, Darshit Vora, Harshil Mehta

## Abstract

Fairness audits of chest radiograph artificial intelligence usually score subgroup performance against labels extracted from radiology reports by natural language processing (NLP). We tested whether these labels can make a model look fairer than it is, a failure we call phantom fairness. On 4,376 NIH ChestX-ray14 images with both NLP and radiologist-adjudicated labels, we audited classifiers from three backbones by sex, age, projection and older women. NLP labels missed 54% to 59% of radiologist-confirmed airspace opacity, pneumothorax and nodule or mass, and every fracture. These misses were not random: models scored them lower than NLP-caught positives for every finding and backbone (median rank-biserial correlation 0.20 for the primary backbone and 0.16 for the others; all adjusted p ≤ 0.038), so the labelling errors hid the models’ own mistakes. Subgroup gaps were larger on adjudicated labels in 16 of 24 comparisons, none individually significant. For pneumothorax in women older than 60 years, the NLP audit showed a sensitivity gap of 0.064 against a true gap of 0.184; removing the hardest positives reproduced this concealment (0.124 against 0.120 observed), whereas random label errors produced less (0.077). Fairness audits based on NLP-extracted labels should be treated as provisional until checked against radiologist-adjudicated labels.

## 1. Introduction

Artificial intelligence (AI) systems that read chest radiographs are among the most mature applications of AI in medical imaging and are moving steadily towards routine clinical use [1]. Before such a system is adopted, hospitals and regulators increasingly ask whether it performs equally well for different groups of patients. Audits of this kind have repeatedly found that chest radiograph classifiers miss disease more often in under-served groups, including women, younger patients and intersectional subgroups [2–6].

Almost every such audit compares the model’s predictions with the labels distributed with the training data. For the most widely used public collection, the National Institutes of Health (NIH) ChestX-ray14 dataset, those labels were not assigned by radiologists. They were extracted automatically from free-text radiology reports by natural language processing (NLP) [7], an approach also used for other large collections such as CheXpert and MIMIC-CXR [8,9]. The reference standard against which fairness is judged is therefore itself imperfect, and label errors of this kind are common in both general and medical image datasets [10–12]. When such errors are concentrated in particular kinds of patients or images, they can hide clinically important failures of a model [13].

The effect on a fairness audit depends on which cases the labels get wrong. The true-positive rate (TPR), or sensitivity, is the proportion of patients with a finding whom the model detects. If an automatic labeller misses positive cases at random, the positives that remain labelled are still representative, and the measured sensitivity is unchanged on average. If instead the labeller tends to miss the same cases that the model also finds hardest, such as subtle findings or poor-quality images, those model errors leave the audit together with their labels. Measured sensitivity is then inflated, and it is inflated most in whichever subgroup loses the most hard cases. Figure 1 shows how this can make a real difference between two subgroups disappear even when the labeller misses positives at the same rate in both. We refer to this apparent equity as phantom fairness.

**Figure 1.**
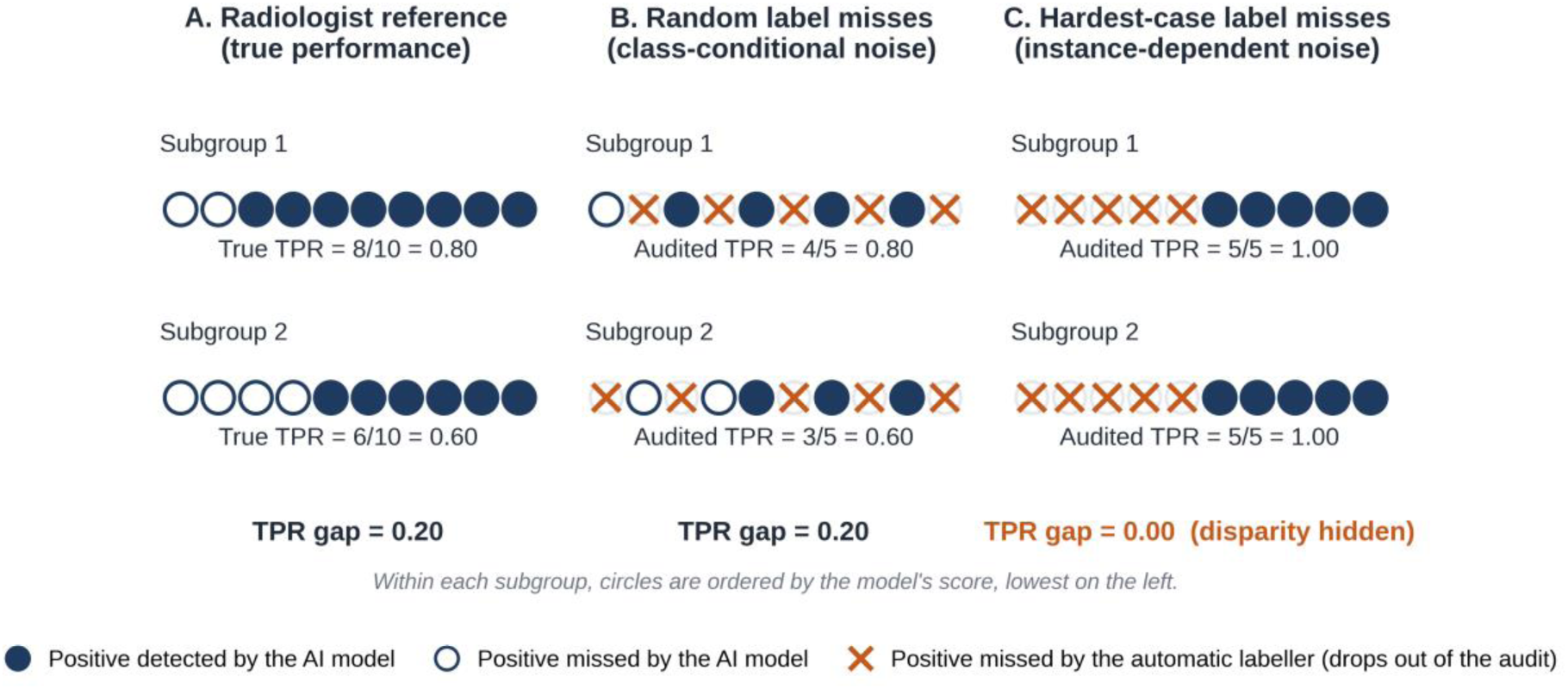
How label errors that fall on the hardest cases can hide a disparity (illustrative example with hypothetical counts). Each circle is a radiograph that radiologists judged positive for a finding; filled circles are detected by the artificial intelligence (AI) model and open circles are missed. (**A**) Against the radiologist reference, the model’s true-positive rate (TPR, sensitivity) is 0.80 in subgroup 1 and 0.60 in subgroup 2, a gap of 0.20. (**B**) An automatic labeller that misses half of the positives at random in each subgroup leaves the audited TPRs, and the gap, unchanged on average. (**C**) A labeller that misses the same proportion of positives, but only those with the lowest model scores, removes the model’s errors from the audit; both subgroups then appear to have a TPR of 1.00 and the disparity is hidden. False-positive labelling errors are omitted for clarity.

The theoretical basis for this concern is established. Wang, Liu and Levy showed that when label-error rates differ between groups, a classifier can satisfy a fairness criterion on noisy labels while violating it on clean labels [14]. Their analysis assumes class-conditional noise, in which errors occur at a fixed rate within each group and class regardless of the individual image. Wu and colleagues extended fair classification to instance-dependent noise, in which the chance of a label error depends on the image itself [15]. Whether report-derived labels in a real imaging dataset behave in an instance-dependent way, and whether this distorts fairness audits, has not been measured. The question matters beyond imaging: a convenient proxy label produced large racial disparities in a widely used health algorithm [16], and imaging models can detect patient characteristics that human readers cannot see [17], which is why responsible and ethical machine learning for health care has become a field in its own right [18,19].

In this study we audited classifiers trained on NIH ChestX-ray14 against radiologist-adjudicated labels for four findings [20], scoring the same images under both label sets so that any change in measured fairness reflects the labels rather than the sample. We asked three questions. First, do subgroup performance gaps differ when measured on NLP labels rather than on adjudicated labels? Second, is the NLP labelling noise instance-dependent, that is, are the positives missed by NLP the cases the model also finds hardest? Third, can label noise of the measured size conceal a real subgroup disparity? To make the audit easy to repeat in other institutions, the whole pipeline runs on a standard laptop processor and every reported number is tied to the code and inputs that produced it.

## 2. Materials and Methods

### 2.1. Data Sources and Label Sets

We used the NIH ChestX-ray14 dataset, which contains 112,120 frontal chest radiographs from 30,805 patients with fourteen finding labels extracted from radiology reports by NLP [7,21]. As the reference standard we used the radiologist-adjudicated labels released by Majkowska and colleagues for four findings (airspace opacity, pneumothorax, nodule or mass, and fracture) on a subset of ChestX-ray14 images [20,22]. We used the version of this label file distributed with the TorchXRayVision library, which covers 4,376 images [23]. The study design is summarised in Figure 2. A 24-image public sample of the PadChest dataset [24,25] was examined only to confirm that the pipeline runs on a second dataset; it was too small for inference and is reported in the Supplementary Materials.

**Figure 2.**
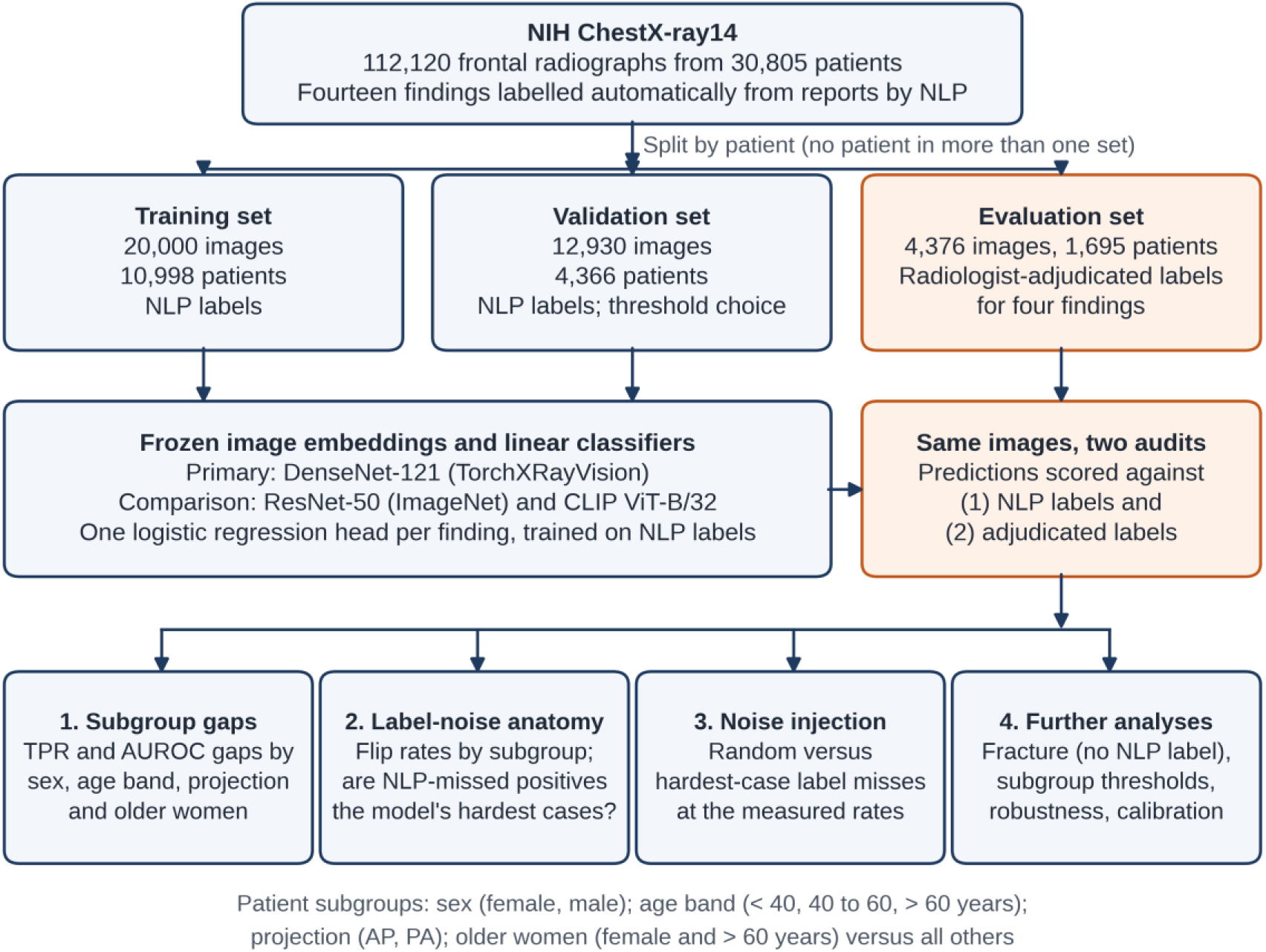
Study design. Patients were split so that no patient contributed images to more than one set. All patients with at least one radiologist-adjudicated image were held out, and their adjudicated images formed the evaluation set, so the adjudicated labels were never used for training or threshold selection. Classifiers were trained on labels extracted by natural language processing (NLP), and the same evaluation images were then audited twice, once against NLP labels and once against adjudicated labels. AP, anteroposterior; AUROC, area under the receiver operating characteristic curve; CLIP, Contrastive Language-Image Pre-training; PA, posteroanterior; TPR, true-positive rate.

### 2.2. Findings and Patient Subgroups

All label sets were mapped to the four findings covered by the adjudicated reference (Table S1). Under the NLP labels, airspace opacity was defined as the union of the infiltration and consolidation labels, nodule or mass as the union of the nodule and mass labels, and pneumothorax by its own label. ChestX-ray14 has no fracture label, so every image was negative for fracture under the NLP labels; this is an extreme form of missing-label noise, which we analysed separately (Section 2.8). Patient age was cleaned by setting values above 120 years to missing.

We examined four subgroup axes, defined in the same way throughout: sex as recorded in the NIH metadata (the field is named ‘Patient Gender’ but records only female or male, so we treat it as recorded sex; gender identity was not available); age band (under 40, 40 to 60, and over 60 years); radiographic projection, posteroanterior (PA) or anteroposterior (AP); and an intersectional axis comparing women older than 60 years with all other patients, referred to as older women. Sex and age were examined because published audits report underdiagnosis in women and younger patients [2,3], and the older-women axis tests one intersection of the two. Projection was included because AP radiographs are usually taken in less mobile or more acutely ill patients and differ in image quality. ChestX-ray14 does not record race or ethnicity, so these could not be studied. Subgroups with fewer than 20 images were excluded, which removed a single evaluation image with missing age. Subgroup sizes are given in Table S2.

### 2.3. Cohort Construction

Splits were made at patient level to prevent leakage. All 26,921 images from the 1,695 patients with at least one adjudicated image were held out, and their 4,376 adjudicated images formed the evaluation set. Of the remaining patients, 15% were assigned at random to a validation set (12,930 images from 4,366 patients), and a random sample of 20,000 images from the rest formed the training set (10,998 patients). The comparison between NLP and adjudicated labels was always made on the same evaluation images, so differences in measured fairness cannot be explained by a change of sample.

### 2.4. Image Embeddings and Classifiers

To isolate the effect of the labels, and to keep the pipeline runnable on a central processing unit (CPU), we used frozen image embeddings with a linear classifier for each finding rather than training networks end to end. The primary backbone was the DenseNet-121 network distributed with TorchXRayVision (weights densenet121-res224-all, 1024-dimensional features) [23,26]. According to its model documentation, this network was pretrained on several chest radiograph collections, including ChestX-ray14, PadChest and the adjudicated NIH labels used here as the reference. To check that our findings did not depend on this exposure, we repeated the key analyses with two backbones that were not trained on the adjudicated labels: an ImageNet-trained ResNet-50 (2048 dimensions) [27] and the Contrastive Language-Image Pre-training (CLIP) ViT-B/32 model with OpenAI weights (512 dimensions) [28]. Images were resized to 224 × 224 pixels, and embeddings were extracted once and cached.

For each backbone, features were standardised using the training set, and one logistic regression classifier per finding (regularisation parameter C = 1.0, class-balanced weights, up to 2,000 iterations) was trained on the NLP labels for airspace opacity, pneumothorax, and nodule or mass. The decision threshold for each finding was set by maximising Youden’s index on the validation set, again against NLP labels [29], giving thresholds of 0.474, 0.084 and 0.537, respectively, for the primary backbone.

This mirrors how a model developed in the usual way would inherit the NLP labels at both training and threshold selection. The adjudicated labels were used only for evaluation.

### 2.5. Fairness Metrics and Statistical Analysis

For each finding and subgroup axis we computed two gaps: the TPR gap at the operating threshold, which corresponds to the equal-opportunity criterion [30], and the gap in the area under the receiver operating characteristic curve (AUROC). A gap was defined as the largest minus the smallest subgroup value on that axis. Confidence intervals (CIs) for individual AUROCs used DeLong variance with the fast midrank algorithm [31,32], and CIs for gaps came from 5,000 bootstrap resamples drawn within subgroups [33].

The main comparison was the regime difference, defined as the gap measured on adjudicated labels minus the gap measured on NLP labels for the same images. A positive value means that an audit based on NLP labels understates the disparity. Its 95% CI and two-sided p value came from a paired bootstrap of evaluation images (5,000 resamples), and p values were adjusted with the Benjamini-Hochberg procedure across the 24 comparisons (three findings, four axes and two metrics) [34]. Because the adjudicated sample gives limited power for individual comparisons, we treated this family as descriptive and the instance-dependence test (Section 2.6) as the primary analysis. This ordering was set before the analyses were run but was not publicly preregistered.

Calibration against adjudicated labels was summarised with the expected calibration error (ECE), using 12 equal-width and 12 equal-mass bins, and with the Brier score, overall and within subgroups [35,36]. We also compared the disparity in the model’s positive-prediction rates with the disparity in adjudicated prevalence, and computed directional bias amplification [37].

### 2.6. Anatomy of the Label Noise

For each subgroup *z* and finding, we estimated the false-negative flip rate of the NLP labels, ε_z_^+^ = P(NLP = 0 | adjudicated = 1), and the false-positive flip rate, ε_z_^−^ = P(NLP = 1 | adjudicated = 0). Disagreement between NLP and adjudicated labels was tested with McNemar’s test [38].

To test whether the noise was instance-dependent, we took the images that radiologists judged positive and compared the model’s scores for those that the NLP labels missed with those that the NLP labels caught. Under class-conditional noise the two groups would have similar score distributions; under instance-dependent noise the missed positives would score lower. We used a one-sided Mann-Whitney U test (caught greater than missed) [39] and summarised the effect as the rank-biserial correlation, *r* = 2*U*/(*n*_1_*n*_2_) − 1, where a positive value means that missed positives score lower [40]. This use of model scores to identify probable label errors follows the principle of confident learning [41]. The analysis was repeated for all three backbones, and p values were adjusted across the nine tests with the Benjamini-Hochberg procedure.

### 2.7. Noise-Injection Experiment

To see whether noise of the measured size could conceal a real disparity, we started from the adjudicated labels of the evaluation images and injected the measured subgroup flip rates in two ways. In the random, class-conditional arm, each positive label was switched to negative with probability ε_z_^+^ and each negative label to positive with probability ε_z_^−^, independently of the model’s score. In the hardest-case, instance-dependent arm, the same number of positives (ε_z_^+^ multiplied by the number of positives in the subgroup, rounded) was removed by taking the positives with the lowest model scores, while false positives were added at random at rate ε_z_^−^ as before. The hardest-case arm therefore represents the limiting situation in which the labeller misses only the hardest positives, the data-generating contrast formalised by Wu and colleagues [15].

For each arm we recomputed the TPR gap at the operating threshold and averaged it over 200 Monte Carlo repetitions. Concealment was defined as the true gap on adjudicated labels minus the audited gap, so that a positive value means that the audit understates the disparity, and it was compared with the concealment actually observed with NLP labels. The experiment covered six cells (three findings by the sex and older-women axes). In the two cells with the largest observed concealment, we also multiplied the flip rates by a factor λ from 0 to 1.5 in 13 steps (100 repetitions per step) to trace a dose-response curve.

### 2.8. Fracture as a Limiting Case

Because ChestX-ray14 contains no fracture label, an NLP-based audit cannot detect any fracture disparity: no image is labelled positive, so the TPR is undefined and the measured gap is zero by construction. To show what such an audit misses, we scored fracture with the primary backbone’s own fracture output and measured subgroup gaps against the adjudicated fracture labels. Because no NLP labels exist from which to set a threshold, the fracture threshold was chosen with Youden’s index on the adjudicated labels of the evaluation set itself, so these results are descriptive.

### 2.9. Threshold-Based Mitigation and Robustness

We tested equal-opportunity post-processing with subgroup-specific thresholds [30]. The evaluation patients were split at random into calibration and test halves 25 times. In each split, a target TPR was set as the overall TPR at a threshold of 0.5 in the calibration half, and a threshold was chosen in each subgroup to reach that target, using either adjudicated or NLP labels in the calibration half. The resulting TPR gap was measured on the test half against adjudicated labels and compared with a single threshold of 0.5, and the cost in accuracy was summarised as balanced accuracy. A cell was analysed only if every subgroup had at least 20 adjudicated positives in the calibration half. For robustness, we repeated the injection experiment with the older-women axis defined by age cut-offs of 55, 60 and 65 years (100 repetitions each).

### 2.10. Reproducibility, Software and Reporting

All analyses were run in three Jupyter notebooks on a Windows laptop without a graphics processing unit, and a fourth notebook generates every table and figure in this article and the Supplementary Materials from the saved results. We used Python 3.11.15, PyTorch 2.12.1 (CPU build), NumPy 2.4.4, pandas 3.0.3 and Matplotlib 3.11.0, together with scikit-learn 1.9.1, SciPy 1.18.1, statsmodels 0.15.1, TorchXRayVision 1.5.5 and open_clip 3.3.0. The random seed was fixed at 1997, every input file was hashed with SHA-256, embeddings were cached under deterministic keys, and a provenance manifest links each reported number to the file that produced it. The code, aggregate result tables and provenance manifests are available at https://github.com/Krutarth007/phantom-fairness. Reporting follows the TRIPOD+AI statement [42], and the completed checklist is provided as Checklist S1. Generative AI tools were used to help debug code, to prepare plotting code for figures from the result tables, and to edit and restructure the manuscript text; all analyses were designed, run and checked by the authors (see Acknowledgments).

## 3. Results

### 3.1. Cohort and Label Agreement

The evaluation set contained 4,376 adjudicated images from 1,695 patients, of which 4,375 entered the subgroup analyses. Most were AP radiographs (3,244 of 4,375, 74.1%), and 361 (8.3%) came from women older than 60 years (Table S2). Finding prevalence differed sharply between label sets (Table 1, Figure S1). On the evaluation images, NLP labelled airspace opacity in 26.5% whereas radiologists identified it in 49.5%, and fracture, which has no NLP label, was present in 4.3%. Agreement between the two label sets ranged from 64.5% for airspace opacity to 95.7% for fracture. The NLP labels missed far more positives than they added for airspace opacity (1,280 false negatives against 275 false positives) and for nodule or mass (349 against 199; both p < 0.001), whereas errors were balanced for pneumothorax (129 against 129; p = 0.95).

**Table 1.** Finding prevalence and agreement between labels extracted by natural language processing (NLP) and radiologist-adjudicated labels.

| Finding | NLP prevalence, full dataset (n = 112,120) | NLP prevalence, evaluation set (n = 4,376) | Adjudicated prevalence, evaluation set (n = 4,376) | Agreement | NLP false negatives | NLP false positives | McNemar p |
| --- | --- | --- | --- | --- | --- | --- | --- |
| Airspace opacity | 20.8% (23,340) | 26.5% (1,161) | 49.5% (2,166) | 64.5% | 1,280 | 275 | < 0.001 |
| Pneumothorax | 4.7% (5,302) | 5.4% (238) | 5.4% (238) | 94.1% | 129 | 129 | 0.95 |
| Nodule or mass | 10.0% (11,207) | 10.4% (455) | 13.8% (605) | 87.5% | 349 | 199 | < 0.001 |
| Fracture <sup>a</sup> | 0.0% (0) | 0.0% (0) | 4.3% (186) | 95.7% | 186 | 0 | < 0.001 |

Prevalence is shown as a percentage with the number of positive images. Agreement, false negatives (adjudicated positive, NLP negative), false positives (adjudicated negative, NLP positive) and McNemar tests (with continuity correction) refer to the 4,376 evaluation images that carry both label sets. <sup>a</sup> ChestX-ray14 has no fracture label, so every adjudicated fracture is an NLP false negative.

### 3.2. Model Performance and Calibration

Measured against adjudicated labels, discrimination was moderate, as expected for linear classifiers on frozen embeddings. Sex-specific AUROCs in women and men were 0.81 and 0.84 for airspace opacity, 0.75 and 0.69 for pneumothorax, and 0.69 and 0.70 for nodule or mass (Figure S2). Calibration was good for airspace opacity (ECE 0.054) but poor for pneumothorax (0.183) and nodule or mass (0.246), reaching 0.292 for nodule or mass in older women (Table S3, Figure S3). Poor calibration for the less common findings is expected after training with class-balanced weights.

### 3.3. Subgroup Gaps on NLP Versus Adjudicated Labels

On identical images, the gap measured on adjudicated labels exceeded the gap measured on NLP labels in 16 of 24 comparisons, with a median regime difference of 0.021 (Table 2, Figure 3). The pattern differed by metric. AUROC gaps were larger on adjudicated labels in 10 of 12 comparisons (median difference 0.029), whereas TPR gaps were larger in only 6 of 12 (median difference 0.001). The largest differences involved the older-women axis. For pneumothorax, the TPR gap was 0.064 on NLP labels and 0.184 on adjudicated labels (difference 0.120, 95% CI −0.044 to 0.236), and for nodule or mass the AUROC gap was 0.034 and 0.153, respectively (difference 0.119, 95% CI −0.007 to 0.211). Only one comparison had an unadjusted p value below 0.05 (AUROC gap for airspace opacity by projection, difference 0.067, 95% CI 0.013 to 0.122, p = 0.018), and none remained significant after Benjamini-Hochberg correction (all adjusted p ≥ 0.43; Table S4).

**Figure 3.**
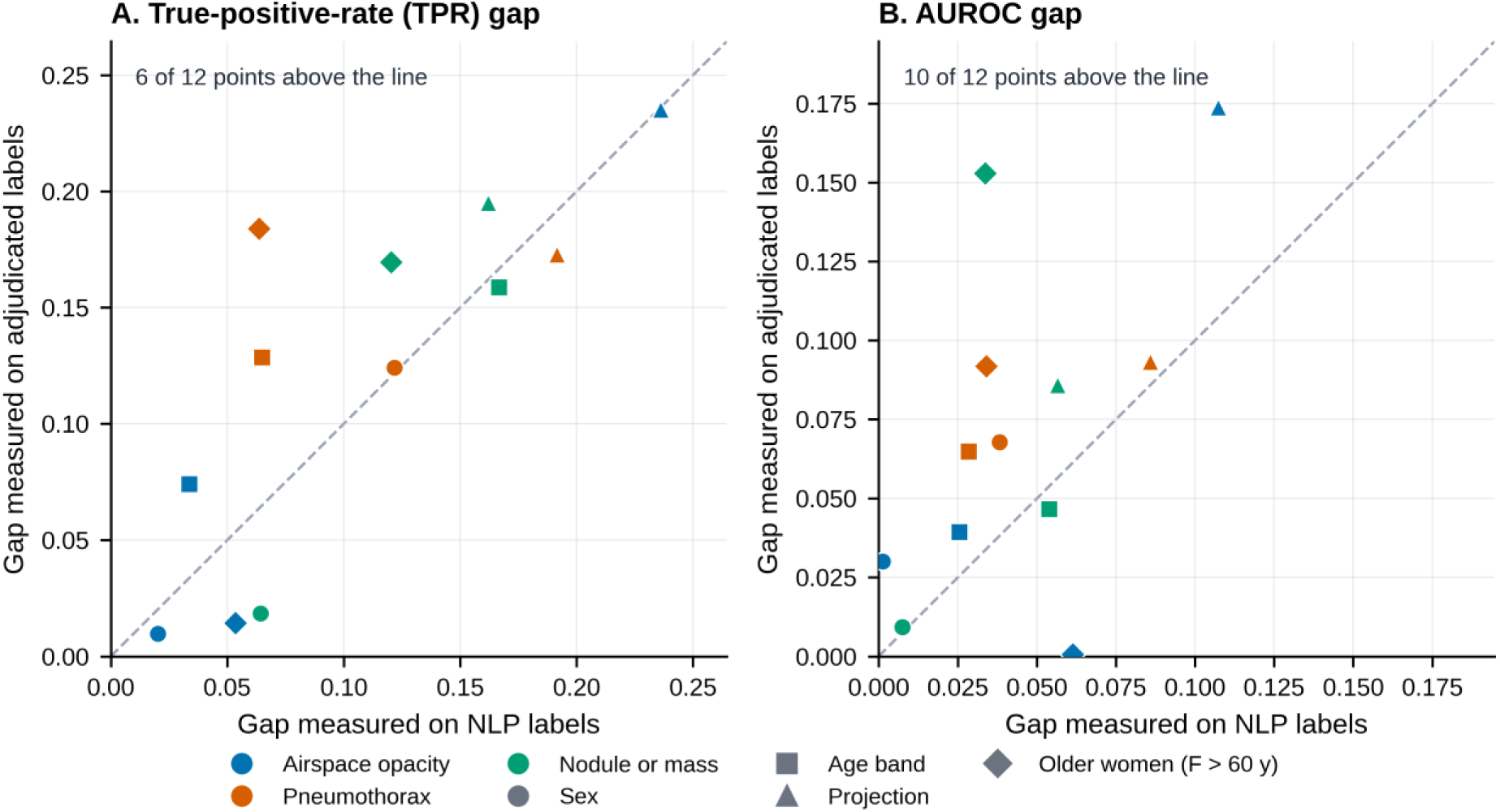
Subgroup gaps measured on adjudicated labels plotted against the same gaps measured on natural language processing (NLP) labels for identical evaluation images. (**A**) True-positive-rate (TPR) gaps at the operating threshold. (**B**) Gaps in the area under the receiver operating characteristic curve (AUROC). Each point is one finding (colour) on one subgroup axis (shape). Points above the dashed line of equality are comparisons in which the NLP-based audit understates the disparity. Confidence intervals are given in Table 2.

**Table 2.** Subgroup gaps measured on NLP labels and on adjudicated labels for the same evaluation images.

| Finding | Axis | TPR gap |  |  | AUROC gap |  |  |
| --- | --- | --- | --- | --- | --- | --- | --- |
|  |  | NLP labels | Adjudicated labels | Difference (95% CI) | NLP labels | Adjudicated labels | Difference (95% CI) |
| Airspace opacity | Sex | 0.020 | 0.010 | −0.010 (−0.051, 0.025) | 0.001 | 0.030 | 0.029 (−0.019, 0.047) |
|  | Age band | 0.034 | 0.074 | 0.040 (−0.043, 0.090) | 0.025 | 0.039 | 0.014 (−0.043, 0.053) |
|  | Projection | 0.236 | 0.235 | −0.001 (−0.078, 0.077) | 0.107 | 0.174 | 0.067 (0.013, 0.122) |
|  | Older women | 0.053 | 0.014 | −0.039 (−0.115, 0.059) | 0.061 | 0.001 | −0.061 (−0.104, 0.020) |
| Pneumothorax | Sex | 0.122 | 0.124 | 0.002 (−0.110, 0.122) | 0.038 | 0.068 | 0.030 (−0.046, 0.097) |
|  | Age band | 0.065 | 0.129 | 0.064 (−0.085, 0.191) | 0.028 | 0.065 | 0.037 (−0.062, 0.105) |
|  | Projection | 0.192 | 0.173 | −0.019 (−0.136, 0.096) | 0.086 | 0.093 | 0.007 (−0.067, 0.084) |
|  | Older women | 0.064 | 0.184 | 0.120 (−0.044, 0.236) | 0.034 | 0.092 | 0.058 (−0.056, 0.136) |
| Nodule or mass | Sex | 0.064 | 0.019 | −0.046 (−0.119, 0.045) | 0.007 | 0.009 | 0.002 (−0.051, 0.043) |
|  | Age band | 0.167 | 0.159 | −0.008 (−0.117, 0.105) | 0.054 | 0.047 | −0.007 (−0.070, 0.060) |
|  | Projection | 0.162 | 0.195 | 0.033 (−0.061, 0.125) | 0.057 | 0.086 | 0.029 (−0.029, 0.085) |
|  | Older women | 0.120 | 0.169 | 0.049 (−0.127, 0.208) | 0.034 | 0.153 | 0.119 (−0.007, 0.211) |

The regime difference was not stable across backbones. For TPR gaps on the sex and older-women axes, it was positive in 3 of 6 cells with DenseNet-121, 5 of 6 with ResNet-50 and 3 of 6 with CLIP, and every interval included zero (Table S5). Measures of bias amplification gave no consistent picture: the difference between prediction and prevalence disparities was positive in 4 of 6 cells, whereas directional bias amplification was negative in 5 of 6 (Table S6). The ordering of the NLP and adjudicated TPR gaps also changed across decision thresholds (Figure S4), so a comparison at one operating point should not be generalised to all thresholds. Taken together, subgroup gaps measured on NLP labels often differed from those measured on adjudicated labels, but the evaluation set was too small to establish the size or direction of any single difference.

### 3.4. The Labelling Noise Was Group-Dependent and Instance-Dependent

False-negative flip rates were high throughout and varied across subgroups, from 0.42 (pneumothorax, PA projection) to 0.76 (airspace opacity, PA projection); excluding projection, they ranged from 0.47 to 0.62 (Figure 4, Table S7). False-positive flip rates were much lower, from 0.02 to 0.14. In other words, the NLP labels missed roughly half or more of the positives that radiologists identified, at rates that differed between subgroups.

**Figure 4.**
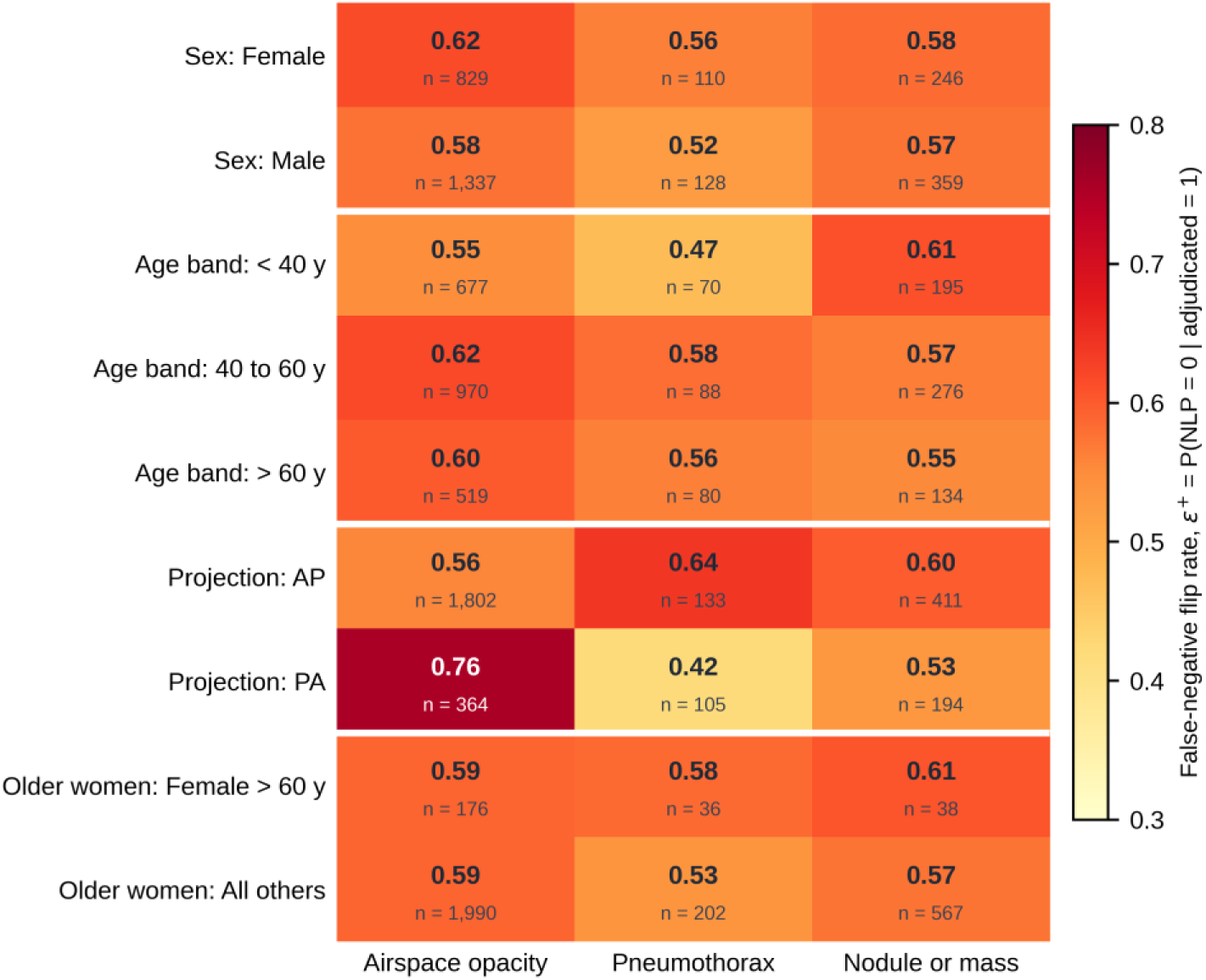
False-negative flip rates of the natural language processing (NLP) labels by subgroup and finding. ε^+^ = P(NLP = 0 | adjudicated = 1) is the probability that a radiograph judged positive by radiologists is labelled negative by NLP. The number below each rate is the number of adjudicated positives in that cell. White horizontal lines separate the four subgroup axes. AP, anteroposterior; PA, posteroanterior; y, years.

The missed positives were not a random sample (Table 3). Among adjudicated positives, the primary model scored NLP-missed cases lower than NLP-caught cases for every finding, with rank-biserial correlations of 0.20 for airspace opacity, 0.17 for pneumothorax and 0.24 for nodule or mass. The same pattern held for ResNet-50 and CLIP, neither of which was trained on the adjudicated labels (median rank-biserial correlation 0.16), and all nine comparisons remained significant after correction (adjusted p ≤ 0.038). The NLP labeller therefore tended to miss the positives that the models also found hardest, which is the instance-dependent noise that the mechanism in Figure 1 requires.

**Table 3.** Instance-dependence of the NLP labelling noise across three image backbones. Among radiographs judged positive by radiologists, model scores are compared between cases that NLP labelling missed and cases that it caught.

| Backbone | Finding | NLP-missed positives (n) | NLP-caught positives (n) | Mean score, missed | Mean score, caught | Rank-biserial r | Adjusted p |
| --- | --- | --- | --- | --- | --- | --- | --- |
| DenseNet-121 (TorchXRayVision) <sup>a</sup> | Airspace opacity | 1,280 | 886 | 0.625 | 0.700 | 0.20 | < 0.001 |
|  | Pneumothorax | 129 | 109 | 0.430 | 0.546 | 0.17 | 0.018 |
|  | Nodule or mass | 349 | 256 | 0.506 | 0.624 | 0.24 | < 0.001 |
| ResNet-50 (ImageNet) | Airspace opacity | 1,280 | 886 | 0.557 | 0.633 | 0.16 | < 0.001 |
|  | Pneumothorax | 129 | 109 | 0.218 | 0.287 | 0.16 | 0.019 |
|  | Nodule or mass | 349 | 256 | 0.357 | 0.453 | 0.17 | < 0.001 |
| CLIP ViT-B/32 | Airspace opacity | 1,280 | 886 | 0.626 | 0.683 | 0.17 | < 0.001 |
|  | Pneumothorax | 129 | 109 | 0.569 | 0.641 | 0.13 | 0.038 |
|  | Nodule or mass | 349 | 256 | 0.481 | 0.541 | 0.15 | 0.001 |

A positive rank-biserial correlation means that NLP-missed positives received lower scores, that is, they were the harder cases for the model. p values are from one-sided Mann-Whitney U tests, adjusted with the Benjamini-Hochberg procedure across the nine tests. <sup>a</sup> Primary backbone; its pretraining data included the adjudicated labels, whereas ResNet-50 and CLIP were not trained on them. CLIP, Contrastive Language-Image Pre-training; NLP, natural language processing.

### 3.5. Noise-Injection Experiment

Table 4 compares the concealment observed with NLP labels with the concealment produced by the two injection arms. Observed concealment was positive in three of the six cells and negative in the other three, where the NLP audit showed a larger gap than the adjudicated labels. The clearest case was pneumothorax in older women: the true TPR gap of 0.184 appeared as 0.064 under NLP labels, a concealment of 0.120. Removing the hardest positives at the measured rates reproduced this closely (0.124), whereas random flips at the same rates produced less (0.077), and in the dose-response analysis only the hardest-case arm reached the observed NLP gap at the measured noise level (Figure S5). In the other cells the two arms were less distinct. For nodule or mass in older women, both arms gave values close to the observed concealment of 0.049 (0.057 for random and 0.054 for hardest-case injection). For pneumothorax by sex, where almost no concealment was observed (0.002), the hardest-case arm overshot (0.081). Averaged over the six cells, the two arms were similarly close to the observed values (mean absolute difference 0.020 for hardest-case and 0.022 for random injection).

**Table 4.**
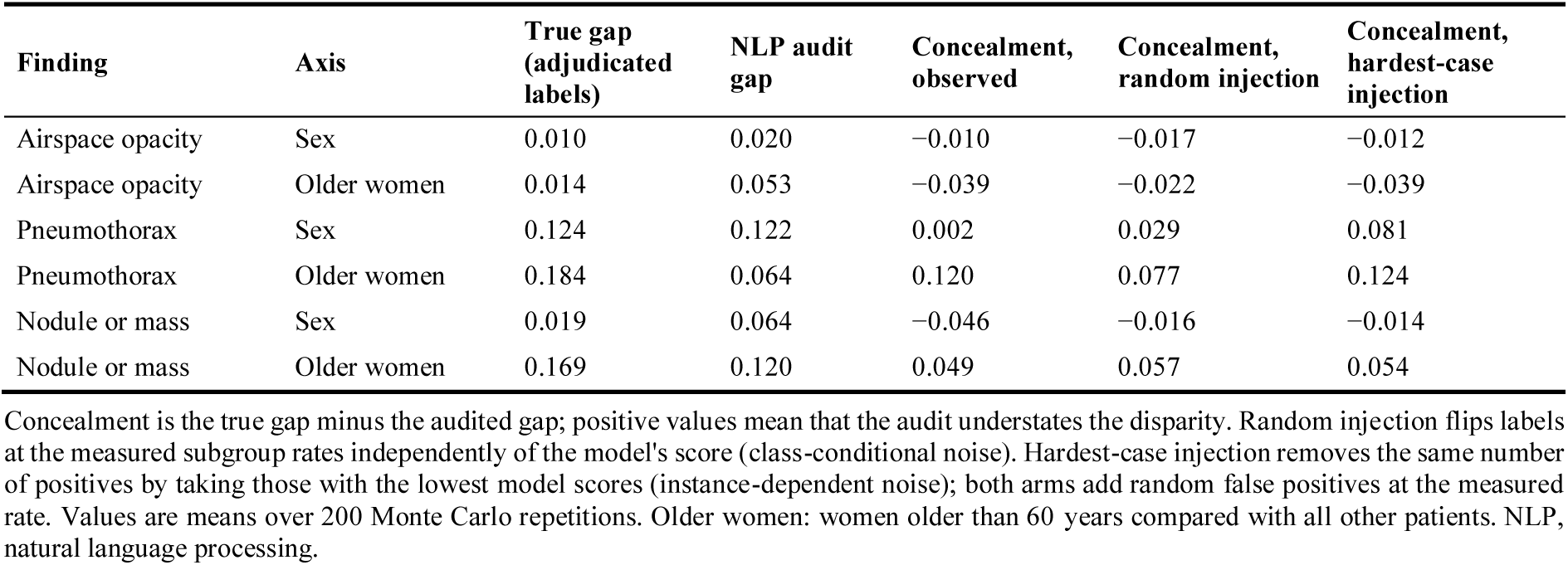
Concealment of the true-positive-rate gap by NLP labels and by injected label noise at the measured subgroup flip rates.

| Finding | Axis | True gap (adjudicated labels) | NLP audit gap | Concealment, observed | Concealment, random injection | Concealment, hardest-case injection |
| --- | --- | --- | --- | --- | --- | --- |
| Airspace opacity | Sex | 0.010 | 0.020 | -0.010 | -0.017 | -0.012 |
| Airspace opacity | Older women | 0.014 | 0.053 | -0.039 | -0.022 | -0.039 |
| Pneumothorax | Sex | 0.124 | 0.122 | 0.002 | 0.029 | 0.081 |
| Pneumothorax | Older women | 0.184 | 0.064 | 0.120 | 0.077 | 0.124 |
| Nodule or mass | Sex | 0.019 | 0.064 | -0.046 | -0.016 | -0.014 |
| Nodule or mass | Older women | 0.169 | 0.120 | 0.049 | 0.057 | 0.054 |

Concealment is the true gap minus the audited gap; positive values mean that the audit understates the disparity. Random injection flips labels at the measured subgroup rates independently of the model's score (class-conditional noise). Hardest-case injection removes the same number of positives by taking those with the lowest model scores (instance-dependent noise); both arms add random false positives at the measured rate. Values are means over 200 Monte Carlo repetitions. Older women: women older than 60 years compared with all other patients. NLP, natural language processing.

Two points help to interpret these results. First, random injection also concealed part of the gap because it adds false positives at subgroup-specific rates. A randomly added positive is usually a true negative that the model does not detect, so it lowers the measured TPR by different amounts in different subgroups, as the group-dependent noise model predicts [14]; random removal of positives alone does not change the expected TPR. Second, the hardest-case arm removes only the lowest-scored positives, whereas the measured noise is only partly instance-dependent (rank-biserial correlations of 0.13 to 0.24), so this arm marks an upper limit rather than a prediction. Across older-women age cut-offs of 55, 60 and 65 years, hardest-case injection produced more concealment than random injection in 5 of 9 cells, including all three pneumothorax cells (Table S8).

### 3.6. Fracture

Fracture shows the limiting case of label noise. ChestX-ray14 has no fracture label, so ε^+^ = 1 in every subgroup and an NLP-based audit reports no disparity at all. Against adjudicated labels, the backbone’s fracture output showed a TPR gap of 0.248 across age bands, with smaller gaps by sex (0.047) and for older women (0.038), and AUROC gaps of 0.040 to 0.087 (Table S9). Because the fracture threshold was chosen on the evaluation labels themselves, these values show what an NLP-based audit cannot see rather than the model’s expected performance.

### 3.7. Threshold-Based Mitigation

Four of the six cells had enough adjudicated positives for the mitigation analysis; the pneumothorax and nodule or mass cells for older women did not (Table S10). With a single threshold of 0.5, the median TPR gap on held-out adjudicated images was 0.046. Subgroup-specific thresholds did not reduce this: the median gap was 0.082 when the thresholds were set on adjudicated labels and 0.103 when they were set on NLP labels (Figure S6). Subgroup thresholds narrowed the gap only for pneumothorax by sex (from 0.192 to 0.131 with adjudicated labels and to 0.122 with NLP labels) and widened it in the other three cells, with balanced accuracy between 0.63 and 0.75. At this sample size, post-processing did not repair the disparities.

### 3.8. Exploratory Second Dataset

The 24-image PadChest sample contained five airspace opacity positives, one nodule or mass positive and no pneumothorax positives. The transferred AUROC for airspace opacity was 0.49 (95% CI 0.24 to 0.74), every subgroup in which power could be estimated was underpowered to detect a TPR gap of 0.15, and the primary backbone had been pretrained on PadChest, so these results were not used for inference (Table S11).

## 4. Discussion

### 4.1. Principal Findings

This audit asked whether demographic fairness measured against automatically extracted labels can be trusted for chest radiograph classifiers. The clearest finding concerns the labels themselves. The NLP labels in ChestX-ray14 missed roughly half or more of the positives that radiologists identified, at rates that varied between subgroups, and the positives they missed were consistently the cases that the models also found hardest. This instance-dependence held for all three findings and all three backbones, including two that were never trained on the adjudicated labels, so it is unlikely to reflect how the primary model was built.

Instance-dependent noise of this kind is the condition under which an audit can hide a disparity, because the model’s errors leave the audit together with their labels (Figure 1). Our data show this happening in at least one clinically relevant comparison: for pneumothorax in women older than 60 years, a true TPR gap of 0.184 appeared as 0.064 in the NLP-based audit, and removing the hardest positives at the measured rates reproduced that concealment. Across all comparisons, gaps measured on adjudicated labels were more often larger than gaps measured on NLP labels, particularly for AUROC, but the evaluation set was too small for any single difference to be statistically significant, and the direction varied by metric, threshold and backbone. We therefore regard the mechanism as demonstrated and its typical size as not yet known.

### 4.2. Comparison with Previous Work

Fairness audits of chest radiograph classifiers have documented underdiagnosis in under-served groups and proposed ways to reduce it [2,3,5,43], almost always using the labels distributed with the data as the reference. Our results support treating that reference standard as part of the audit rather than as a given, in line with frameworks for the medical algorithmic audit and with reviews of fairness in computational medicine [44,45]. Wang, Liu and Levy proved that group-dependent label noise can make a classifier appear fair on noisy labels [14], and Wu and colleagues extended fair classification to instance-dependent noise [15]. To our knowledge, this study is among the first to measure instance-dependent labelling noise directly in a widely used clinical imaging dataset and to link it to concealment in a fairness audit. The findings are also consistent with causal accounts that place much of the bias in medical imaging in the data-generating process rather than in the model alone [46,47].

### 4.3. Implications for Practice

Four practical points follow. First, a fairness report computed on automatically extracted labels should be treated as provisional. Hospitals evaluating a chest radiograph AI product, and the companies that supply such products, should ask which labels were used to measure subgroup performance and whether a radiologist-adjudicated reference was included. Second, a modest adjudicated reference set is needed to detect a hidden disparity, and repairing one appears to need more. In our data, subgroup-specific thresholds did not reduce the gaps even when they were set on adjudicated labels, and a disparity that cannot be seen in the audit labels cannot be targeted at all. Third, calibration deserves attention alongside error rates. Calibration was poor for the less common findings in our models, calibration and equal error rates cannot generally be achieved together when prevalence differs between subgroups [48–50], and subgroup calibration should be compared with sample size in mind because the ECE itself depends on subgroup size [51]. Fourth, the audit is inexpensive to repeat. It runs end to end on a laptop processor using frozen embeddings, with every input hashed and every random seed fixed, so an institution can re-run it on its own data whenever a model or labelling pipeline changes. This matters increasingly as chest radiograph classifiers approach radiologist-level reported performance in some tasks [52].

### 4.4. Limitations

This study has several limitations. The adjudicated reference covers four findings and 4,376 images, which limited the power of individual subgroup comparisons and left some intersectional cells too small for the mitigation analysis. Bootstrap intervals and the instance-dependence tests treated images as independent, although many patients contributed several images; intervals that account for clustering within patients would probably be wider. We used linear classifiers on frozen embeddings to isolate the effect of the labels; this strengthens internal validity, but fully fine-tuned networks may interact with label noise differently. The primary backbone had been pretrained on data that included the adjudicated labels and PadChest, which we addressed by repeating the key analyses with two backbones that had not seen those labels. The injection experiment compared two simplified forms of noise, and the real noise lies between them. The fracture analysis used a threshold chosen on the evaluation labels and is descriptive. Sex was available only as a binary field, and race and ethnicity were not recorded, so important dimensions of fairness could not be examined. Finally, all images came from one institution and one NLP labelling pipeline, so the size of the effect in other datasets and with other labellers is unknown.

### 4.5. Future Research

Larger adjudicated reference sets are needed to estimate how often, and by how much, report-derived labels conceal disparities, and the instance-dependence test should be applied to other datasets, labelling pipelines and imaging modalities [53]. Because the test needs only model scores and a small set of trusted labels, it could also guide which images are sent for expert review: the positives that a model scores lowest are those most likely to be mislabelled and most likely to hide a disparity.

## 5. Conclusions

In NIH ChestX-ray14, labels extracted automatically from radiology reports missed a large share of true positives, and the positives they missed were the cases that classifiers also found hardest. Label noise of this kind can make a model look fairer than it is, and in one intersectional comparison it hid most of a real disparity. Fairness reports built on automatically extracted labels should therefore be regarded as provisional, and a small radiologist-adjudicated reference set should be part of any fairness audit of chest radiograph AI. The reproducible, laptop-based audit described here offers one practical way to provide it.

## Supplementary Materials

Table S1: Mapping of label vocabularies to the four findings; Table S2: Subgroup sizes in the full dataset and in the evaluation set; Figure S1: Finding prevalence under NLP and adjudicated labels; Figure S2: Receiver operating characteristic and precision-recall curves by sex against adjudicated labels; Table S3: Calibration overall and by subgroup; Figure S3: Reliability diagram for airspace opacity by sex; Table S4: Full results for the 24 regime-difference comparisons; Table S5: Regime difference by image backbone; Table S6: Bias amplification; Figure S4: True-positive-rate gaps across decision thresholds for airspace opacity by sex; Table S7: Flip rates and McNemar tests by subgroup; Figure S5: Dose-response analysis of the noise-injection experiment; Table S8: Robustness of the noise-injection experiment to the older-women age cut-off; Table S9: Fracture as a limiting case; Table S10: Threshold-based mitigation; Figure S6: Threshold-based mitigation; Table S11: Exploratory PadChest analysis; Checklist S1: TRIPOD+AI checklist.

## Author Contributions

K.P. conceived the study, designed the methodology, wrote the code, performed the analyses and wrote the manuscript. P.B., D.V. and H.M. contributed to the study concept and methodology and critically revised the manuscript. All authors read and approved the final version.

## Institutional Review Board Statement

Ethical review and approval were waived for this study because it used only publicly available, fully de-identified images and labels. Under the United States Common Rule (45 CFR 46.102(e)(1)), analysis of non-identifiable data does not constitute human subjects research.

## Informed Consent Statement

Patient consent was waived because only publicly available, de-identified data, released by the data providers under their terms of use, were analysed.

## Data Availability Statement

The data analysed in this study are third-party, de-identified datasets derived from public resources. NIH ChestX-ray14 images and metadata are openly available from the NIH Clinical Center at https://nihcc.app.box.com/v/ChestXray-NIHCC [21]. The radiologist-adjudicated labels of Majkowska and colleagues are available from Google Cloud at https://docs.cloud.google.com/healthcare-api/docs/resources/public-datasets/nih-chest [22] and are distributed with the TorchXRayVision library [23].

The PadChest sample is available from BIMCV at https://bimcv.cipf.es/bimcv-projects/padchest/ under its data use agreement [25]. The analysis code, aggregate result tables and provenance manifests are openly available at https://github.com/Krutarth007/phantom-fairness. No patient images or image-level derived data are shared.

## Acknowledgments

The authors thank the NIH Clinical Center for releasing ChestX-ray14, Majkowska and colleagues for releasing the radiologist-adjudicated labels, and BIMCV for the PadChest dataset.

## Abbreviations

The following abbreviations are used in this manuscript:

AI: artificial intelligence
AP: anteroposterior
AUROC: area under the receiver operating characteristic curve
CI: confidence interval
CLIP: Contrastive Language-Image Pre-training
CPU: central processing unit
ECE: expected calibration error
NIH: National Institutes of Health
NLP: natural language processing
PA: posteroanterior
TPR: true-positive rate
TRIPOD+AI: Transparent Reporting of a multivariable prediction model for Individual Prognosis Or Diagnosis, updated for artificial intelligence

