## Supplementary Materials for "Phantom Fairness: A Reproducible Audit of How Automatically Extracted Labels Can Conceal Demographic Disparities in Chest Radiograph Classifiers"

#### Notes

All values were produced by the analysis notebooks and are tied by SHA-256 hash to the result files in the provenance manifest of the code repository. Older women denotes women older than 60 years compared with all other patients. Gaps are the largest minus the smallest subgroup value on an axis. Abbreviations are defined in the Notes at the start of this file.

Reproducibility details. Embeddings were extracted at 224 × 224 pixels in batches of 32 using 13 CPU threads, without L2 normalisation, and cached under keys built from a SHA-256 digest of the ordered image identifiers. The global random seed was 1997. Bootstrap analyses used 5,000 resamples (2,500 for the backbone comparison in Table S5), the noise-injection experiment used 200 Monte Carlo repetitions (100 for the dose-response and robustness analyses), and the mitigation analysis used 25 random patient-level splits.

**Table S1.** Mapping of the three label vocabularies to the four findings covered by the adjudicated reference.

| Finding | NIH NLP labels | Adjudicated label | PadChest terms (exploratory sample only) |
|---|---|---|---|
| Airspace opacity | Infiltration; Consolidation | Airspace opacity | infiltrates; alveolar pattern; consolidation; pneumonia; air bronchogram; increased density; atypical pneumonia; interstitial pattern; ground glass pattern |
| Pneumothorax | Pneumothorax | Pneumothorax | pneumothorax; hydropneumothorax |
| Nodule or mass | Nodule; Mass | Nodule or mass | nodule; multiple nodules; mass; pulmonary mass; lung mass; pseudonodule |
| Fracture | None (no fracture label in ChestX-ray14) | Fracture | fracture; rib fracture; clavicle fracture; humeral fracture; vertebral fracture; callus rib fracture |

An image is positive for a finding under the NLP labels if any of the listed NLP labels is positive.

**Table S2.** Subgroup sizes in the full NIH ChestX-ray14 dataset and in the evaluation set, with the number of adjudicated positives per finding in the evaluation set.

| Axis | Subgroup | Full dataset (images) | Evaluation set (images) | Airspace opacity positives | Pneumothorax positives | Nodule or mass positives |
|---|---|---|---|---|---|---|
| Sex | Female | 48,780 | 1,765 | 829 | 110 | 246 |
| | Male | 63,340 | 2,610 | 1,337 | 128 | 359 |
| Age band | < 40 y | 35,935 | 1,435 | 677 | 70 | 195 |
| | 40 to 60 y | 49,137 | 1,917 | 970 | 88 | 276 |
| | > 60 y | 27,032 | 1,023 | 519 | 80 | 134 |
| | Missing age | 16 | 1 (excluded) | | | |
| Projection | AP | 44,810 | 3,244 | 1,802 | 133 | 411 |
| | PA | 67,310 | 1,131 | 364 | 105 | 194 |
| Older women | Women > 60 y | 9,798 | 361 | 176 | 36 | 38 |
| | All others | 102,322 | 4,014 | 1,990 | 202 | 567 |

One evaluation image with missing age fell in a subgroup of fewer than 20 images and was excluded, leaving 4,375 images for subgroup analyses. Abbreviations are defined in the Notes at the start of this file.

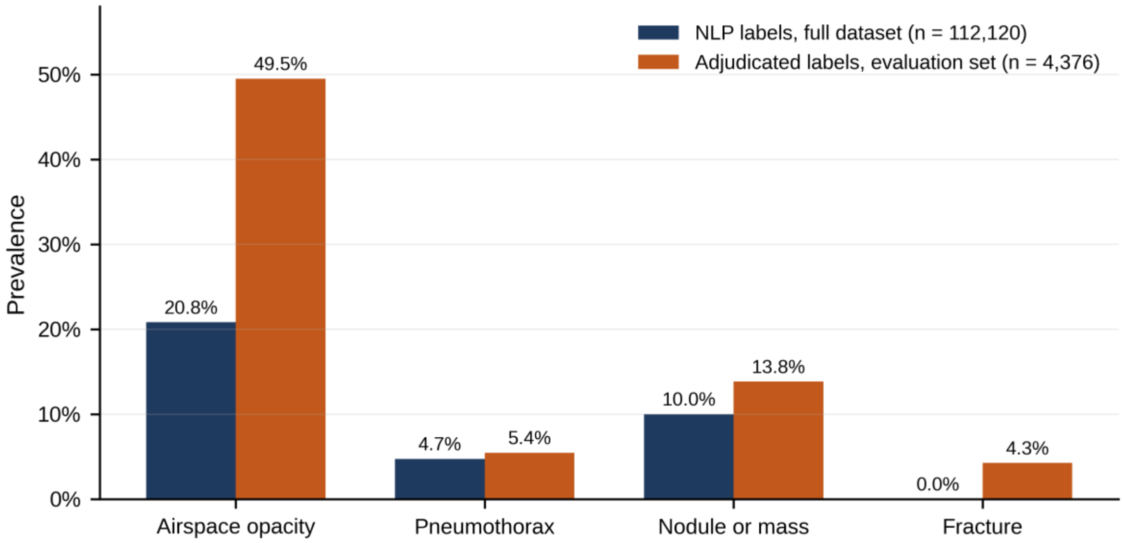


**Figure S1.** Prevalence of the four findings under NLP labels in the full dataset and under radiologist-adjudicated labels in the evaluation set. ChestX-ray14 has no fracture label, so NLP prevalence of fracture is zero by construction.

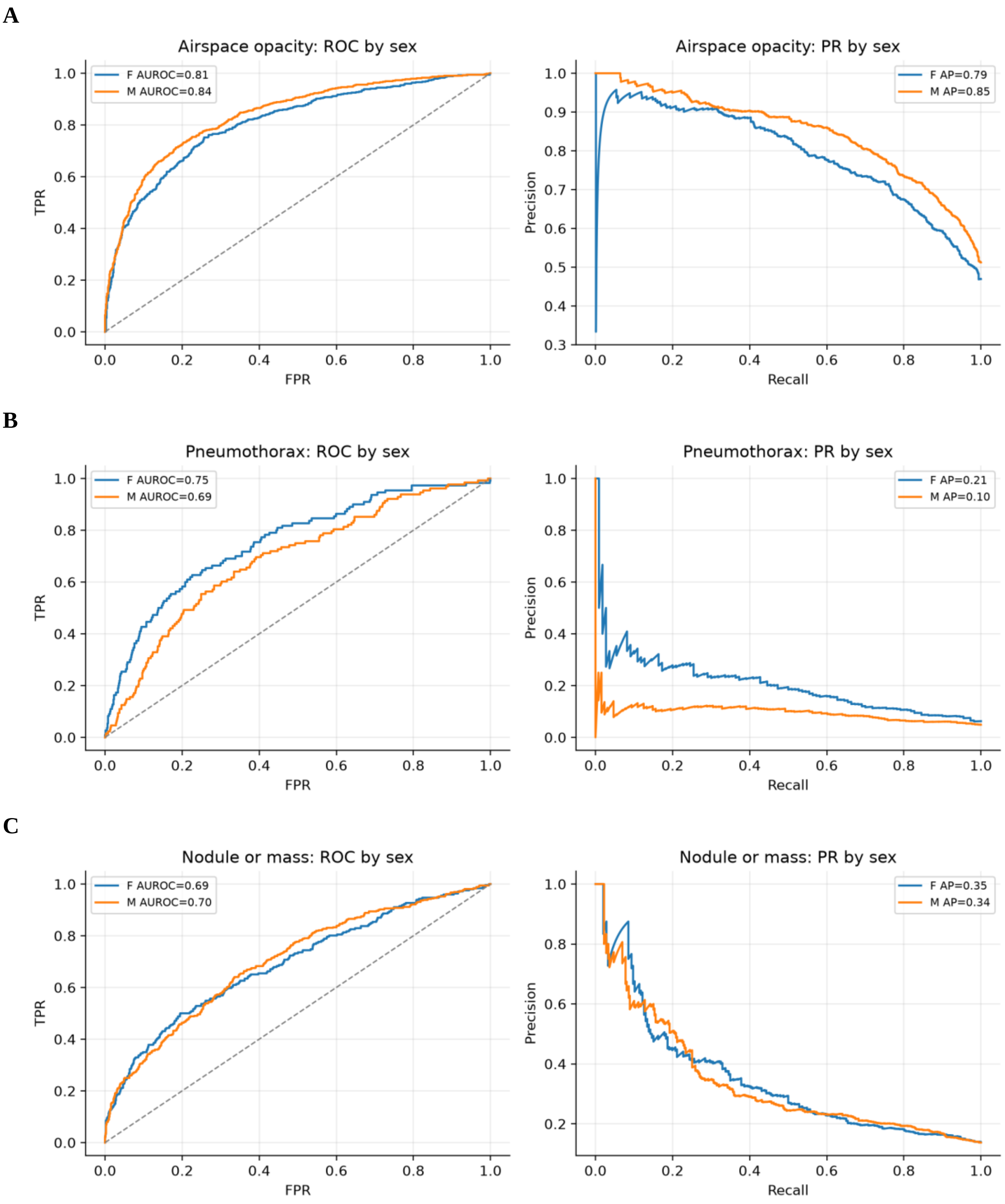


**Figure S2.** Receiver operating characteristic (left) and precision-recall (right) curves by sex for the primary backbone, scored against adjudicated labels: (**A**) airspace opacity; (**B**) pneumothorax; (**C**) nodule or mass. The legend gives the AUROC or average precision (AP in these panels) for female (F) and male (M) patients. FPR, false-positive rate; TPR, true-positive rate.

**Table S3.** Calibration of the primary backbone against adjudicated labels, overall and by subgroup.

| **Finding** | **Axis** | **Subgroup** | **n** | **ECE, equal-width bins** | **ECE, equal-mass bins** | **Brier score** |
|---|---|---|---|---|---|---|
| Airspace opacity | Overall | All | 4,375 | 0.054 | 0.052 | 0.173 |
| | Sex | Female | 1,765 | 0.062 | 0.059 | 0.182 |
| | Sex | Male | 2,610 | 0.054 | 0.053 | 0.167 |
| | Age band | 40 to 60 y | 1,917 | 0.058 | 0.063 | 0.167 |

| Finding | Axis | Subgroup | n | ECE, equal-width bins | ECE, equal-mass bins | Brier score |
|---|---|---|---|---|---|---|
| | Age band | < 40 y | 1,435 | 0.064 | 0.057 | 0.174 |
| | Age band | > 60 y | 1,023 | 0.051 | 0.060 | 0.184 |
| | Older women | Women > 60 y | 361 | 0.062 | 0.058 | 0.173 |
| | Older women | All others | 4,014 | 0.056 | 0.054 | 0.173 |
| Pneumothorax | Overall | All | 4,375 | 0.183 | 0.190 | 0.151 |
| | Sex | Female | 1,765 | 0.203 | 0.210 | 0.165 |
| | Sex | Male | 2,610 | 0.169 | 0.175 | 0.141 |
| | Age band | 40 to 60 y | 1,917 | 0.191 | 0.199 | 0.156 |
| | Age band | < 40 y | 1,435 | 0.175 | 0.182 | 0.140 |
| | Age band | > 60 y | 1,023 | 0.178 | 0.184 | 0.157 |
| | Older women | Women > 60 y | 361 | 0.175 | 0.188 | 0.162 |
| | Older women | All others | 4,014 | 0.184 | 0.190 | 0.150 |
| Nodule or mass | Overall | All | 4,375 | 0.246 | 0.249 | 0.203 |
| | Sex | Female | 1,765 | 0.238 | 0.241 | 0.200 |
| | Sex | Male | 2,610 | 0.252 | 0.253 | 0.205 |
| | Age band | 40 to 60 y | 1,917 | 0.253 | 0.255 | 0.205 |
| | Age band | < 40 y | 1,435 | 0.217 | 0.218 | 0.192 |
| | Age band | > 60 y | 1,023 | 0.277 | 0.277 | 0.214 |
| | Older women | Women > 60 y | 361 | 0.292 | 0.291 | 0.226 |
| | Older women | All others | 4,014 | 0.242 | 0.245 | 0.201 |

ECE used 12 bins. Abbreviations are defined in the Notes at the start of this file.

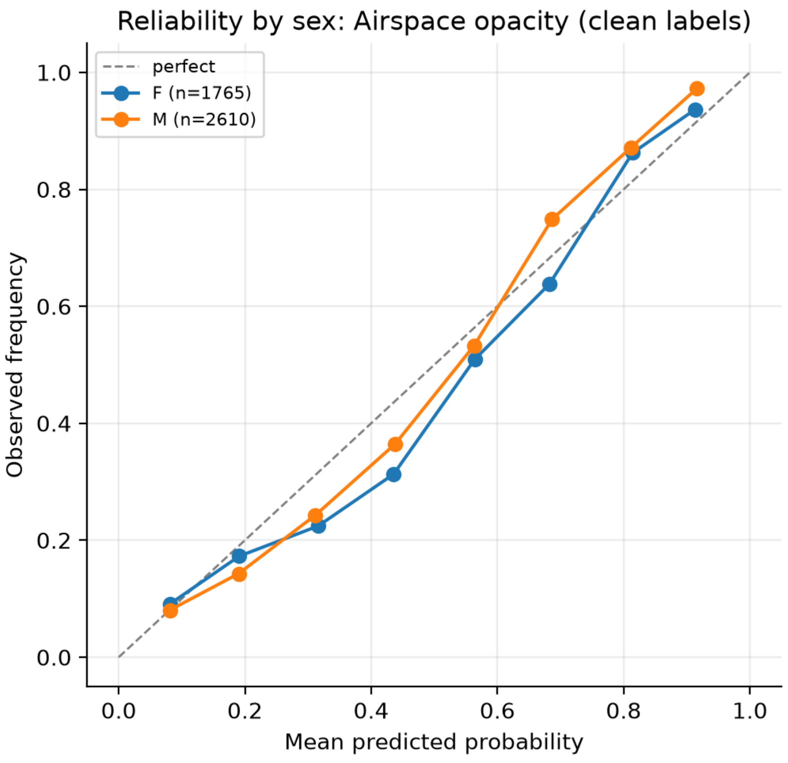


**Figure S3.** Reliability diagram for airspace opacity by sex against adjudicated labels (primary backbone). The dashed line indicates perfect calibration; n is the number of images per group.

**Table S4.** Full results for the 24 comparisons of subgroup gaps measured on NLP labels and on adjudicated labels for the same evaluation images.

| Finding | Axis | Metric | Gap, NLP labels (95% CI) | Gap, adjudicated labels (95% CI) | Difference (95% CI) | p | Adjusted p |
|---|---|---|---|---|---|---|---|
| Airspace opacity | Sex | TPR | 0.020 (0.001, 0.070) | 0.010 (0.001, 0.044) | −0.010 (−0.051, 0.025) | 0.656 | 0.983 |
| | Sex | AUROC | 0.001 (0.001, 0.040) | 0.030 (0.006, 0.055) | 0.029 (−0.019, 0.047) | 0.330 | 0.983 |
| | Age band | TPR | 0.034 (0.009, 0.099) | 0.074 (0.033, 0.119) | 0.040 (−0.043, 0.090) | 0.429 | 0.983 |
| | Age band | AUROC | 0.025 (0.006, 0.072) | 0.039 (0.013, 0.073) | 0.014 (−0.043, 0.053) | 0.750 | 0.992 |
| | Projection | TPR | 0.236 (0.160, 0.311) | 0.235 (0.183, 0.289) | −0.001 (−0.078, 0.077) | 0.992 | 0.992 |
| | Projection | AUROC | 0.107 (0.060, 0.154) | 0.174 (0.138, 0.209) | 0.067 (0.013, 0.122) | 0.018 | 0.432 |
| | Older women | TPR | 0.053 (0.003, 0.133) | 0.014 (0.001, 0.080) | −0.039 (−0.115, 0.059) | 0.534 | 0.983 |
| | Older women | AUROC | 0.061 (0.007, 0.117) | 0.001 (0.001, 0.050) | −0.061 (−0.104, 0.020) | 0.184 | 0.983 |
| Pneumothorax | Sex | TPR | 0.122 (0.024, 0.228) | 0.124 (0.021, 0.229) | 0.002 (−0.110, 0.122) | 0.949 | 0.992 |
| | Sex | AUROC | 0.038 (0.002, 0.106) | 0.068 (0.007, 0.136) | 0.030 (−0.046, 0.097) | 0.476 | 0.983 |
| | Age band | TPR | 0.065 (0.018, 0.210) | 0.129 (0.038, 0.281) | 0.064 (−0.085, 0.191) | 0.436 | 0.983 |
| | Age band | AUROC | 0.028 (0.011, 0.127) | 0.065 (0.016, 0.154) | 0.037 (−0.062, 0.105) | 0.630 | 0.983 |
| | Projection | TPR | 0.192 (0.087, 0.302) | 0.173 (0.068, 0.275) | −0.019 (−0.136, 0.096) | 0.770 | 0.992 |
| | Projection | AUROC | 0.086 (0.020, 0.156) | 0.093 (0.026, 0.161) | 0.007 (−0.067, 0.084) | 0.826 | 0.992 |
| | Older women | TPR | 0.064 (0.004, 0.201) | 0.184 (0.070, 0.282) | 0.120 (−0.044, 0.236) | 0.169 | 0.983 |
| | Older women | AUROC | 0.034 (0.002, 0.126) | 0.092 (0.015, 0.168) | 0.058 (−0.056, 0.136) | 0.384 | 0.983 |
| Nodule or mass | Sex | TPR | 0.064 (0.004, 0.157) | 0.019 (0.002, 0.102) | −0.046 (−0.119, 0.045) | 0.465 | 0.983 |
| | Sex | AUROC | 0.007 (0.001, 0.064) | 0.009 (0.001, 0.058) | 0.002 (−0.051, 0.043) | 0.970 | 0.992 |
| | Age band | TPR | 0.167 (0.063, 0.286) | 0.159 (0.075, 0.269) | −0.008 (−0.117, 0.105) | 0.929 | 0.992 |
| | Age band | AUROC | 0.054 (0.012, 0.126) | 0.047 (0.014, 0.112) | −0.007 (−0.070, 0.060) | 0.910 | 0.992 |
| | Projection | TPR | 0.162 (0.067, 0.258) | 0.195 (0.111, 0.275) | 0.033 (−0.061, 0.125) | 0.505 | 0.983 |
| | Projection | AUROC | 0.057 (0.006, 0.111) | 0.086 (0.038, 0.133) | 0.029 (−0.029, 0.085) | 0.342 | 0.983 |
| | Older women | TPR | 0.120 (0.008, 0.294) | 0.169 (0.023, 0.329) | 0.049 (−0.127, 0.208) | 0.638 | 0.983 |
| | Older women | AUROC | 0.034 (0.002, 0.139) | 0.153 (0.046, 0.266) | 0.119 (−0.007, 0.211) | 0.064 | 0.768 |

Gap intervals are from 5,000 bootstrap resamples within subgroups. The difference (adjudicated gap minus NLP gap), its interval and the two-sided p value are from a paired bootstrap of images (5,000 resamples); adjusted p values use the Benjamini-Hochberg procedure across the 24 comparisons. Abbreviations are defined in the Notes at the start of this file.

**Table S5.** Difference between TPR gaps measured on adjudicated and on NLP labels, by image backbone.

| Backbone | Finding | Axis | Difference (95% CI) | p |
|---|---|---|---|---|
| DenseNet-121 (TorchXRayVision) | Airspace opacity | Sex | −0.010 (−0.050, 0.025) | 0.651 |
| | Airspace opacity | Older women | −0.039 (−0.114, 0.057) | 0.510 |
| | Pneumothorax | Sex | 0.002 (−0.111, 0.125) | 0.946 |
| | Pneumothorax | Older women | 0.120 (−0.047, 0.239) | 0.146 |
| | Nodule or mass | Sex | −0.046 (−0.119, 0.047) | 0.483 |
| | Nodule or mass | Older women | 0.049 (−0.128, 0.206) | 0.618 |
| ResNet-50 (ImageNet) | Airspace opacity | Sex | 0.018 (−0.035, 0.057) | 0.594 |
| | Airspace opacity | Older women | −0.017 (−0.095, 0.046) | 0.697 |
| | Pneumothorax | Sex | 0.023 (−0.106, 0.150) | 0.711 |
| | Pneumothorax | Older women | 0.065 (−0.121, 0.229) | 0.545 |
| | Nodule or mass | Sex | 0.038 (−0.053, 0.114) | 0.420 |
| | Nodule or mass | Older women | 0.122 (−0.101, 0.249) | 0.390 |
| CLIP ViT-B/32 | Airspace opacity | Sex | −0.023 (−0.059, 0.018) | 0.379 |
| | Airspace opacity | Older women | −0.011 (−0.082, 0.039) | 0.636 |

| Backbone | Finding | Axis | Difference (95% CI) | p |
|---|---|---|---|---|
| | Pneumothorax | Sex | 0.018 (−0.094, 0.104) | 0.929 |
| | Pneumothorax | Older women | 0.013 (−0.148, 0.166) | 0.878 |
| | Nodule or mass | Sex | 0.002 (−0.081, 0.087) | 0.955 |
| | Nodule or mass | Older women | −0.058 (−0.225, 0.153) | 0.718 |

Positive values mean that the NLP-based audit understates the TPR gap. Intervals and two-sided p values are from a paired bootstrap of images (2,500 resamples). Each backbone has its own logistic heads and thresholds. Abbreviations are defined in the Notes at the start of this file.

**Table S6.** Bias amplification by the primary model relative to adjudicated labels.

| Finding | Axis | Prevalence disparity | Prediction disparity | Amplification | Directional bias amplification |
|---|---|---|---|---|---|
| Airspace opacity | Sex | 0.043 | 0.010 | −0.032 | −0.016 |
| Airspace opacity | Older women | 0.008 | 0.032 | 0.024 | 0.003 |
| Pneumothorax | Sex | 0.013 | 0.091 | 0.078 | −0.011 |
| Pneumothorax | Older women | 0.049 | 0.079 | 0.030 | −0.056 |
| Nodule or mass | Sex | 0.002 | 0.025 | 0.023 | −0.024 |
| Nodule or mass | Older women | 0.036 | 0.023 | −0.013 | −0.014 |

Prevalence disparity is the difference in adjudicated prevalence between subgroups; prediction disparity is the difference in the proportion predicted positive at the operating threshold; amplification is prediction disparity minus prevalence disparity. Directional bias amplification follows Wang and Russakovsky. The two measures disagree in sign for most cells, so no conclusion about amplification is drawn. Abbreviations are defined in the Notes at the start of this file.

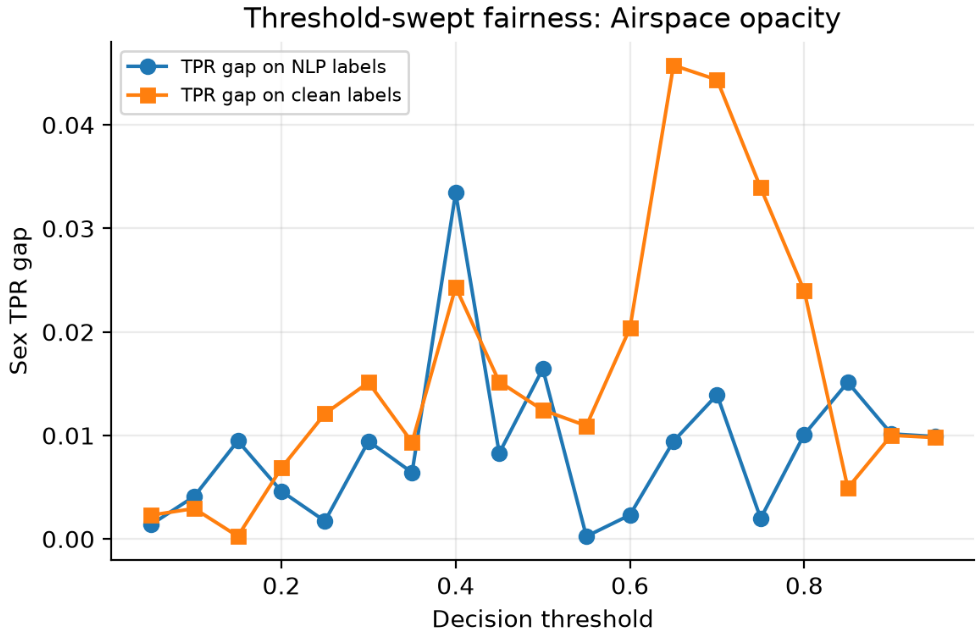


**Figure S4.** TPR gap between female and male patients for airspace opacity across decision thresholds, measured on NLP labels and on adjudicated labels for the same evaluation images (primary backbone). The ordering of the two gaps changes across thresholds, so a comparison at one operating point does not generalise to all thresholds. TPR, true-positive rate.

**Table S7.** Flip rates of the NLP labels against adjudicated labels and model true-positive rates under each label set, by finding and subgroup (primary backbone).

| Finding | Axis | Subgroup | n | Positives (adjudicated) | $\varepsilon^{+}$ | $\varepsilon^{-}$ | NLP-missed positives | McNemar p | TPR, adjudicated | TPR, NLP |
|---|---|---|---|---|---|---|---|---|---|---|
| Airspace opacity | Sex | Female | 1,765 | 829 | 0.62 | 0.120 | 510 | < 0.001 | 0.79 | 0.76 |
| | | Male | 2,610 | 1,337 | 0.58 | 0.128 | 770 | < 0.001 | 0.80 | 0.78 |

| Finding | Axis | Subgroup | n | Positives (adjudicated) | $\varepsilon^+$ | $\varepsilon^-$ | NLP-missed positives | McNemar p | TPR, adjudicated | TPR, NLP |
|---|---|---|---|---|---|---|---|---|---|---|
| | Age band | 40 to 60 y | 1,917 | 970 | 0.62 | 0.127 | 598 | < 0.001 | 0.82 | 0.78 |
| | | < 40 y | 1,435 | 677 | 0.55 | 0.142 | 371 | < 0.001 | 0.80 | 0.76 |
| | | > 60 y | 1,023 | 519 | 0.60 | 0.093 | 311 | < 0.001 | 0.75 | 0.79 |
| | Projection | AP | 3,244 | 1,802 | 0.56 | 0.126 | 1,005 | < 0.001 | 0.84 | 0.81 |
| | | PA | 1,131 | 364 | 0.76 | 0.121 | 275 | < 0.001 | 0.60 | 0.58 |
| | Older women | Women > 60 y | 361 | 176 | 0.59 | 0.076 | 104 | < 0.001 | 0.78 | 0.83 |
| | | All others | 4,014 | 1,990 | 0.59 | 0.129 | 1,176 | < 0.001 | 0.80 | 0.77 |
| Pneumothorax | Sex | Female | 1,765 | 110 | 0.56 | 0.037 | 62 | 1.000 | 0.83 | 0.83 |
| | | Male | 2,610 | 128 | 0.52 | 0.027 | 67 | 1.000 | 0.70 | 0.71 |
| | Age band | 40 to 60 y | 1,917 | 88 | 0.58 | 0.027 | 51 | 1.000 | 0.80 | 0.80 |
| | | < 40 y | 1,435 | 70 | 0.47 | 0.026 | 33 | 0.810 | 0.67 | 0.74 |
| | | > 60 y | 1,023 | 80 | 0.56 | 0.046 | 45 | 0.915 | 0.80 | 0.76 |
| | Projection | AP | 3,244 | 133 | 0.64 | 0.020 | 85 | 0.084 | 0.68 | 0.67 |
| | | PA | 1,131 | 105 | 0.42 | 0.064 | 44 | 0.045 | 0.86 | 0.86 |
| | Older women | Women > 60 y | 361 | 36 | 0.58 | 0.058 | 21 | 0.874 | 0.92 | 0.82 |
| | | All others | 4,014 | 202 | 0.53 | 0.029 | 108 | 0.946 | 0.73 | 0.76 |
| Nodule or mass | Sex | Female | 1,765 | 246 | 0.58 | 0.051 | 143 | < 0.001 | 0.52 | 0.47 |
| | | Male | 2,610 | 359 | 0.57 | 0.054 | 206 | < 0.001 | 0.53 | 0.53 |
| | Age band | 40 to 60 y | 1,917 | 276 | 0.57 | 0.048 | 156 | < 0.001 | 0.57 | 0.53 |
| | | < 40 y | 1,435 | 195 | 0.61 | 0.056 | 119 | < 0.001 | 0.43 | 0.41 |
| | | > 60 y | 1,023 | 134 | 0.55 | 0.058 | 74 | 0.061 | 0.59 | 0.58 |
| | Projection | AP | 3,244 | 411 | 0.60 | 0.050 | 246 | < 0.001 | 0.46 | 0.45 |
| | | PA | 1,131 | 194 | 0.53 | 0.061 | 103 | < 0.001 | 0.66 | 0.61 |
| | Older women | Women > 60 y | 361 | 38 | 0.61 | 0.056 | 23 | 0.532 | 0.37 | 0.39 |
| | | All others | 4,014 | 567 | 0.57 | 0.053 | 326 | < 0.001 | 0.54 | 0.51 |

$\varepsilon^+ = P(NLP = 0 \mid adjudicated = 1)$; $\varepsilon^- = P(NLP = 1 \mid adjudicated = 0)$. McNemar tests compare NLP and adjudicated labels within each subgroup (continuity-corrected, or exact when fewer than 25 discordant pairs). TPRs are at the operating threshold. Abbreviations are defined in the Notes at the start of this file.

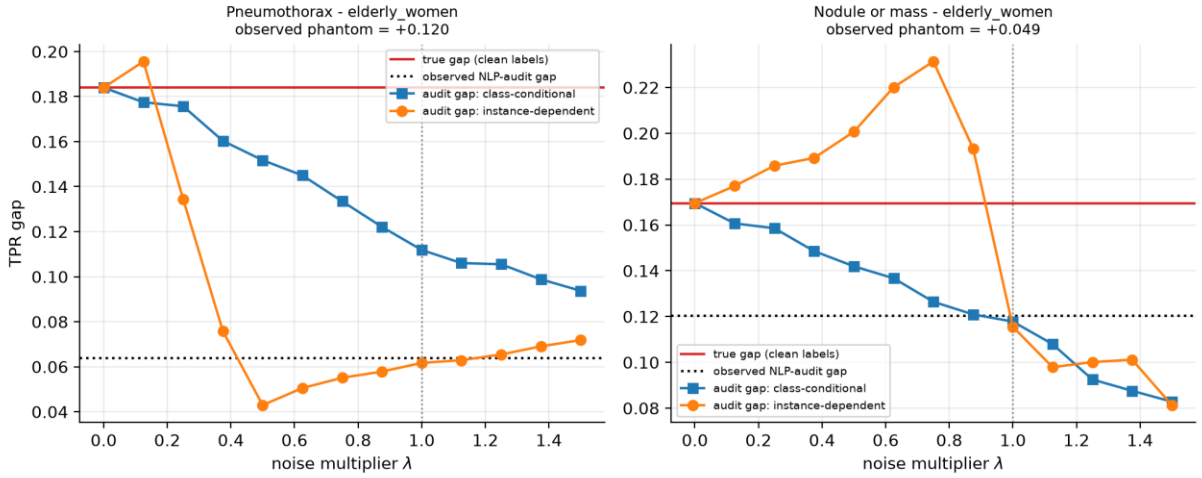


**Figure S5.** Dose-response analysis of the noise-injection experiment in the two cells with the largest observed concealment: pneumothorax (left) and nodule or mass (right), each for women older than 60 years compared with all other patients (labelled elderly_women in the panels). The measured subgroup flip rates were multiplied by λ (λ = 1 is the measured level). The red line is the true TPR gap on adjudicated labels and the dotted black line is the gap observed with NLP labels. Blue squares show random (class-conditional) injection and orange circles show hardest-case (instance-dependent) injection; each point is the mean of 100 Monte Carlo repetitions. TPR, true-positive rate.

**Table S8.** Robustness of the noise-injection experiment to the age cut-off used to define older women.

| Finding | Age cut-off for older women | Concealment, observed | Concealment, random injection | Concealment, hardest-case injection | Hardest-case greater than random |
|---|---|---|---|---|---|
| Airspace opacity | > 55 y | 0.013 | −0.016 | −0.011 | Yes |
| Pneumothorax | > 55 y | 0.075 | 0.043 | 0.085 | Yes |
| Nodule or mass | > 55 y | −0.130 | −0.014 | −0.023 | No |
| Airspace opacity | > 60 y | −0.039 | −0.021 | −0.040 | No |
| Pneumothorax | > 60 y | 0.120 | 0.072 | 0.122 | Yes |
| Nodule or mass | > 60 y | 0.049 | 0.052 | 0.054 | Yes |
| Airspace opacity | > 65 y | −0.035 | −0.050 | −0.060 | No |
| Pneumothorax | > 65 y | 0.025 | 0.046 | 0.058 | Yes |
| Nodule or mass | > 65 y | 0.061 | 0.003 | −0.061 | No |

Concealment is the true TPR gap minus the audited gap; positive values mean that the audit understates the disparity. Means over 100 Monte Carlo repetitions. TPR, true-positive rate; y, years.

**Table S9.** Fracture as a limiting case of label noise: subgroup gaps visible to an NLP-based audit and against adjudicated labels.

| Axis | Gap visible to an NLP-based audit | TPR gap, adjudicated labels | AUROC gap, adjudicated labels | Difference in adjudicated prevalence |
|---|---|---|---|---|
| Sex | 0.000 | 0.047 | 0.040 | 0.030 |
| Age band | 0.000 | 0.248 | 0.087 | 0.061 |
| Older women | 0.000 | 0.038 | 0.075 | 0.022 |

ChestX-ray14 has no fracture label, so no image is NLP-positive and an NLP-based audit can show no disparity. Fracture scores came from the primary backbone's own fracture output, and the threshold was chosen with Youden's index on the adjudicated labels of the evaluation set, so the values are descriptive. Abbreviations are defined in the Notes at the start of this file.

**Table S10.** Equal-opportunity mitigation with subgroup-specific thresholds: TPR gaps on held-out adjudicated images.

| Finding | Axis | Single threshold (0.5) | Subgroup thresholds, adjudicated labels | Subgroup thresholds, NLP labels | Balanced accuracy | Splits used | Status |
|---|---|---|---|---|---|---|---|
| Airspace opacity | Sex | 0.029 | 0.052 | 0.041 | 0.746 | 25 | Analysed |
| Airspace opacity | Older women | 0.031 | 0.041 | 0.117 | 0.749 | 25 | Analysed |
| Pneumothorax | Sex | 0.192 | 0.131 | 0.122 | 0.650 | 25 | Analysed |
| Pneumothorax | Older women | | | | | 0 | Too few positives |
| Nodule or mass | Sex | 0.060 | 0.111 | 0.089 | 0.633 | 25 | Analysed |
| Nodule or mass | Older women | | | | | 0 | Too few positives |

Means over 25 random patient-level splits into calibration and test halves. A cell was analysed only if every subgroup had at least 20 adjudicated positives in the calibration half. Balanced accuracy refers to thresholds set on adjudicated labels. Abbreviations are defined in the Notes at the start of this file.

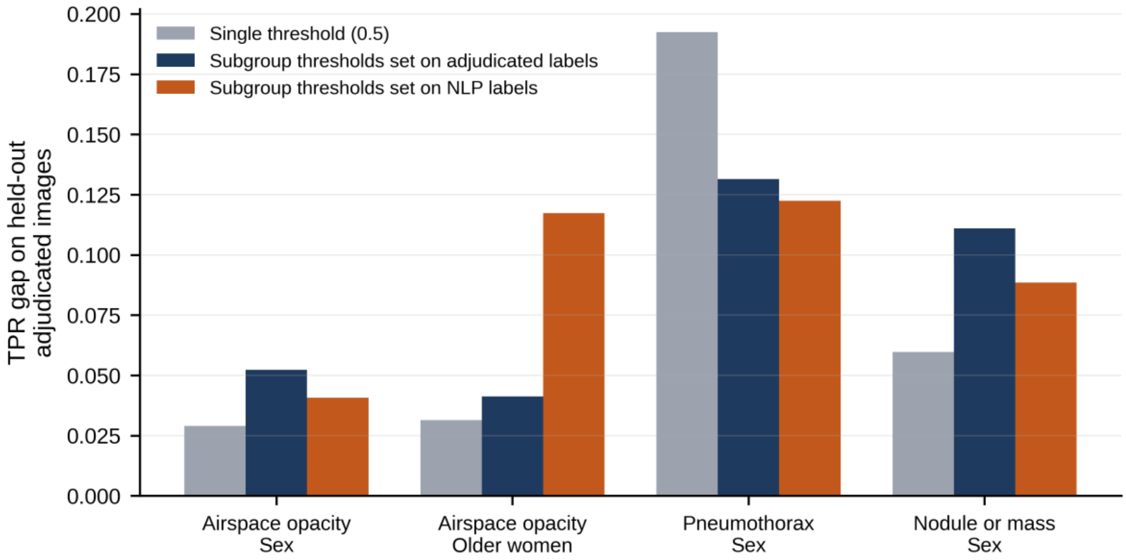


**Figure S6.** TPR gaps on held-out adjudicated images with a single threshold of 0.5 and with subgroup-specific thresholds set on adjudicated or on NLP labels, for the four cells with enough adjudicated positives. Values are means over 25 random patient-level splits. NLP, natural language processing; TPR, true-positive rate.

**Table S11.** Exploratory PadChest analysis (24-image public sample). The primary backbone was pretrained on PadChest, so these results are not independent of the model and were not used for inference. The sample contained five airspace opacity positives, no pneumothorax positives, one nodule or mass positive and two fracture positives. The transferred AUROC for airspace opacity was 0.49 (95% CI 0.24, 0.74); with a single positive, the AUROC for nodule or mass is not interpretable. The table gives subgroup sizes and the minimum TPR gap detectable in each subgroup.

| Axis | Subgroup | n | Minimum detectable TPR gap | Underpowered for a gap of 0.15 |
|---|---|---|---|---|
| Sex | Male | 13 | 0.55 | Yes |
| Sex | Female | 11 | 0.60 | Yes |
| Age band | > 60 y | 11 | 0.60 | Yes |
| Age band | < 40 y | 8 | 0.70 | Yes |
| Age band | 40 to 60 y | 5 | 0.89 | Yes |
| Projection | PA | 14 | 0.53 | Yes |
| Projection | Other | 9 | 0.66 | Yes |
| Projection | AP | 1 | | Not estimable |

Abbreviations are defined in the Notes at the start of this file.

### Checklist S1. TRIPOD+AI Checklist

Phantom Fairness: A Reproducible Audit of How Automatically Extracted Labels Can Conceal Demographic Disparities in Chest Radiograph Classifiers

TRIPOD+AI is the reporting guideline for studies that develop or evaluate prediction models, including those built with machine learning [Collins GS et al. *BMJ* 2024, 385, e078378]. This study audits chest radiograph classifiers: it trains simple logistic classifiers on frozen image embeddings and evaluates them against two sets of labels, with the aim of testing how label noise affects fairness audits. It does not propose a model for clinical use, so items about implementation and user interaction are marked N/A with the reason. Locations refer to the numbered sections, tables and figures of the manuscript. The wording of each item is abbreviated; the full text is in the published guideline.

| Item | Topic and checklist item | Reported | Where reported | Comment |
|---|---|---|---|---|
| | **Title** | | | |
| 1 | Identify the study as developing or evaluating a prediction model, the target population and the outcome. | Yes | Title | The title names the study as an audit of chest radiograph classifiers and the labelling issue examined. |
| | **Abstract** | | | |
| 2 | Structured summary of the study design, setting, methods, results and conclusions. | Yes | Abstract | Single unstructured paragraph of 200 words, as the journal requires; it covers data, methods, results and conclusions. |
| | **Introduction** | | | |
| 3a | Health care context, rationale, and intended use of the model. | Yes | Section 1 | Fairness audits of chest radiograph AI before clinical adoption. |
| 3b | Target population and intended purpose in the care pathway, including intended users. | Yes | Sections 1, 4.3 | Users of the audit are hospitals, regulators and suppliers evaluating chest radiograph AI. |
| 3c | Known health inequalities between sociodemographic groups. | Yes | Section 1 | Underdiagnosis in women, younger patients and intersectional subgroups [2–6]. |
| 4 | Study objectives. | Yes | Section 1 (final paragraph) | Three questions stated. |
| | **Methods** | | | |
| 5a | Sources of data for development and evaluation, rationale, and representativeness. | Yes | Sections 2.1, 2.3; Figure 2; Table S2; Section 4.4 | Public NIH ChestX-ray14 with NLP labels for development; adjudicated subset for evaluation. Single-institution source acknowledged. |
| 5b | Dates of data collection. | No | None | Secondary public datasets; acquisition dates are given by the data providers [7,21]. |
| 6a | Study setting and number and location of centres. | Yes | Sections 2.1, 4.4 | One institution (NIH Clinical Center). |
| 6b | Eligibility criteria for participants. | Yes | Sections 2.1 to 2.3 | All frontal radiographs; the evaluation set is the adjudicated subset; subgroups under 20 images excluded. |
| 6c | Treatments received and how they were handled. | N/A | None | Diagnostic imaging study; treatment is not relevant to the outcome. |
| 7 | Data preprocessing and quality checking, including across sociodemographic groups. | Yes | Sections 2.2, 2.4 | Age values above 120 years set to missing; images resized to 224 × 224 and features standardised; the same pipeline for all subgroups. |
| 8a | Outcome definition, time horizon, how and when assessed, and consistency across groups. | Yes | Sections 2.1, 2.2, 2.6; Table S1 | Presence of four findings on the radiograph (no time horizon). Consistency of labelling across groups is the main question of the study. |
| 8b | Qualifications and demographics of outcome assessors when interpretation is subjective. | Partly | Section 2.1 | Adjudication by radiologists [20]; assessor demographics are not available. |
| 8c | Blinding of outcome assessment. | N/A | None | Labels were produced by the data providers before this study. |

| Item | Topic and checklist item | Reported | Where reported | Comment |
|---|---|---|---|---|
| 9a | Choice of initial predictors. | Yes | Section 2.4 | Frozen image embeddings from three backbones; no clinical predictors. |
| 9b | How predictors were measured. | Yes | Section 2.4 | Embedding extraction at 224 × 224 pixels, once, cached. |
| 9c | User interaction in measuring predictors. | N/A | None | Fully automated; no user interaction. |
| 10 | How the study size was arrived at. | Partly | Sections 2.3, 2.5, 4.4 | Size was set by the available adjudicated images; no formal calculation. Limited power is discussed. |
| 11 | Handling of missing data. | Yes | Sections 2.2, 3.1 | One evaluation image with missing age was excluded from subgroup analyses. |
| 12a | How the data were used and partitioned. | Yes | Section 2.3; Figure 2 | Patient-level split into training, validation and evaluation sets. |
| 12b | How predictors were handled. | Yes | Section 2.4 | Standardised using the training set. |
| 12c | Model type, building steps, hyperparameters and internal validation. | Yes | Sections 2.4, 2.9 | Logistic regression (C = 1.0, balanced class weights, 2,000 iterations); no hyperparameter search; patient-disjoint hold-out evaluation. |
| 12d | Heterogeneity across clusters. | Partly | Sections 2.5, 4.4 | Backbone comparison reported (Table S5). Clustering of images within patients was not modelled; this is a stated limitation. |
| 12e | Performance measures and plots, with rationale. | Yes | Sections 2.5, 2.6 | TPR and AUROC gaps, calibration, flip rates, rank-biserial correlation; bootstrap intervals. |
| 12f | Model updating, overall or for particular groups. | Yes | Section 2.9 | Subgroup-specific thresholds tested as post-processing. |
| 12g | How predictions were calculated for evaluation. | Yes | Sections 2.4, 2.10 | Code in the public repository. |
| 13 | Methods for class imbalance. | Yes | Section 2.4 | Class-balanced weights; prevalence in Table 1. |
| 14 | Approaches to address model fairness, with rationale. | Yes | Sections 2.5, 2.9 | Equal-opportunity gaps, bias amplification and subgroup thresholds. |
| 15 | Model output and how thresholds were chosen. | Yes | Section 2.4 | Probabilities; thresholds 0.474, 0.084 and 0.537 from Youden's index on the validation set. |
| 16 | Differences between development and evaluation data. | Yes | Sections 2.3, 3.1; Table 1; Figure S1 | Development used NLP labels; evaluation used adjudicated labels on patient-disjoint images; prevalence differences reported. |
| 17 | Ethics committee approval and consent procedure. | Yes | Institutional Review Board and Informed Consent statements | Not required: public, de-identified data. |
| 18a | Funding. | Yes | Funding statement | No external funding. |
| 18b | Conflicts of interest. | Yes | Conflicts of Interest statement | |
| 18c | Protocol availability. | Partly | Section 2.5 | No protocol; the order of analyses was set before they were run but was not preregistered. |
| 18d | Registration. | N/A | None | Not registered; the study is a secondary analysis of public data. |
| 18e | Data sharing. | Yes | Data Availability Statement | Sources and links given; no patient images are redistributed. |
| 18f | Code sharing. | Yes | Section 2.10; Data Availability Statement | Three notebooks plus a fourth that regenerates all tables and figures; result tables and provenance hashes. |
| 19 | Patient and public involvement. | N/A | None | There was no patient or public involvement; the study used public secondary data. |
| | **Results** | | | |
| 20a | Flow of participants through the study. | Yes | Section 2.3; Figure 2; Section 3.1 | Images and patients in each set reported. |
| 20b | Characteristics of participants, including key demographics, outcome events and missing data. | Yes | Table 1; Table S2; Section 3.1 | Sex, age band, projection and older-women subgroup sizes with the number of positives. Race and ethnicity are not recorded in the dataset. |

| Item | Topic and checklist item | Reported | Where reported | Comment |
|---|---|---|---|---|
| 20c | Comparison of development and evaluation data distributions. | Yes | Table 1; Table S2; Figure S1 | Prevalence and subgroup sizes in the full dataset and evaluation set. |
| 21 | Number of participants and outcome events in each analysis. | Yes | Section 3.1; Tables 1, 3; Table S2 | |
| 22 | Full model specification to allow predictions and third-party evaluation. | Partly | Sections 2.4, 2.10 | Settings and code to retrain the classifiers are provided. The classifiers are research tools for the audit and are not proposed for clinical use. |
| 23a | Model performance with confidence intervals, including key subgroups. | Yes | Tables 2, 3, 4; Tables S3 to S7; Figures 3, 4 | |
| 23b | Heterogeneity in performance across clusters. | Partly | Table S5; Section 3.3 | Examined across backbones and thresholds (Figure S4); not across centres, because there is one source. |
| 24 | Results of any model updating. | Yes | Section 3.7; Table S10; Figure S6 | |
| | **Discussion** | | | |
| 25 | Overall interpretation, including fairness, in the context of objectives and previous work. | Yes | Sections 4.1, 4.2 | |
| 26 | Limitations and their effects on bias, uncertainty and generalisability. | Yes | Section 4.4 | |
| 27a | Handling of poor-quality or unavailable input data when implementing the model. | N/A | None | The study evaluates an audit method and proposes no deployed model. |
| 27b | Whether users must interact with input data, and expertise required. | N/A | None | No user interaction. |
| 27c | Next steps for research, with a view to applicability and generalisability. | Yes | Section 4.5 | |
